# Generalizable electroencephalographic classification of Parkinson’s Disease using deep learning

**DOI:** 10.1101/2022.08.30.22279401

**Authors:** Richard James Sugden, Phedias Diamandis

**Affiliations:** Department of Medical Biophysics, University of Toronto, Toronto, Ontario, M5S 1A8, Canada; Department of Laboratory Medicine and Pathobiology, University of Toronto, Toronto, Ontario, M5S 1A8, Canada; Princess Margaret Cancer Center, University Health Network, Toronto, Ontario, 610 University Avenue, M5G 2C1, Canada; Laboratory Medicine Program, University Health Network, 200 Elizabeth Street, Toronto, Ontario, M5G 2C4, Canada

**Keywords:** Electroencephalography, Deep Learning, Parkinson’s Disease, Classification, Convolutional Neural Network

## Abstract

There is growing interest in using electroencephalography (EEG) and deep learning (DL) to aid in the diagnosis of neurological conditions like Parkinson’s Disease (PD). Many existing DL approaches to classify PD from EEG data cite performance metrics in the high 90% accuracies, but may be grossly overestimating their real-word capabilities due to information-leakage between training and testing data. Our aim was to characterize the potential of deep learning for classifying PD using a conservative training approach with unseen external testing data. We used publicly available resting-state EEG data from patients with PD from two seperate centers (University of New Mexico (n = 54) and University of Iowa (n = 28)) for our training and testing sets, respectively. We implemented a channelwise convolutional neural network and tuned it using a subjectwise cross validation approach. We found that an approach commonly cited in the literature overestimated performance in excess of 20%, while our pipeline more conservatively estimated performance by epoch (accuracy: 69.2%; sensitivity: 66.5%; specificity: 72.2%) and by subject (accuracy: 77.4%, sensitivity: 76.9%, specificity: 77.8%). Moreover, we show that our model generalized well to an unseen and external testing dataset without degradation in performance by epoch (accuracy: 77.2; sensitivity: 83.5%; specificity: 71.0%) and by subject (accuracy: 83.8%, sensitivity: 88.6%, specificity: 79.0%). These results highlight the effect of information leakage and serve as a new benchmark for future generalization of DL approaches to classify PD using EEG data.

## Introduction

Electroencephalography (EEG) is a neuroimaging technique where electrodes are placed on the subject’s scalp to detect voltages originating from local neural activity. This non-invasive technology has long been used in the diagnosis of diseases like epilepsy, sleep disorders, sensory transmission [1]. In these diseases, EEG changes are fairly marked, allowing expert neurophysiologists to make subjective interpretations on underlying disease states. Other neurological conditions may also show changes, but the complexity of EEG signals may preclude reliable assessment by human observers or existing tools. More recently, advancements in deep-learning (DL) and neural networks have prompted enthusiasm and investigations to determine whether these modern computational techniques can enable EEG to be used to supplement the diagnosis of other common and complex diseases such as Alzheimer’s disease, brain tumors, and Parkinson’s disease (PD) [2, 3].

Specifically, PD is a neurological condition associated with symptoms such as tremors, rigidity, akinesia, and postural instability with a prevalence of roughly 1% of the population [4]. These somewhat subjective clinical criteria lie along a spectrum and do not present uniformly between patients and disease progression, which motivates the search for ancillary and objective biomarkers of the disease. Recently, several papers have used various pre-processing and machine learning techniques to demonstrate the potential for EEG to be used as a biomarker for PD. Many of these studies report very high accuracies above 98%. [5 - 12] However, the methods employed in these articles may suffer from the limitation of information-leakage between the training and testing data [6]. This limitation can lead to large overestimation of classification ability on unseen, real-world data. Such marked decreases in testing and real world performance can create confusion in the literature and decrease enthusiasm of these tools within the community at large. To the best of our knowledge, no research has reported the true potential of DL model classification performance on unseen PD subjects using an external dataset. Here, to address this important gap, we present a carefully constructed convolutional neural network (CNN) based pipeline to classify PD from resting-state EEG data and demonstrate its ability to generalize.

## Methods

### Cohort Development

For our training and test datasets, we used publicly available data from the University of New Mexico (UNM) and University of Iowa (UI) respectively. Both datasets were collected from patients with PD (UNM: N=54, PD=27; UI: N=28, PD=14) who had not taken levodopa medication in the 12 hours prior to data collection. Data was collected in the resting condition with the eyes open. The EEG hardware was the 64-channel Brain Vision system with sintered Ag/AgCl electrodes collected over the range of 0.1–100 Hz with a sampling rate of 500 Hz. Data was accessed on PRED+CT (www.predictsite.com) further details about the cohorts and collection protocols are described by [13].

### Preprocessing

To match the UNM data, the UI data was re-referenced to channel CPz (International 10-20 system) as a monopolar reference. Electrodes were removed if they were not placed at the same location in both datasets, resulting in 60 selected non-reference EEG channels. Data was bandpass filtered between 1-45Hz using default settings of the filter provided in the MNE-Python package. Each data was segmented into uniform non-overlapping epochs representing five seconds (2500 data points per channel) and was normalized to a mean of zero and standard deviation of one.

### Machine Learning

Using PyTorch, we implemented a channelwise 1D CNN with an appended feed-forward network classifier adapted from Oh et al. and adjusted to account for the different input data shape [4]. Using a batch size of 8 for 30 epochs of training, the CNN was trained by optimization of cross-entropy loss using an Adam optimizer with a learning rate of 0.0001.

### Classification

To estimate classification capability from the training data, we used a leave-one-out cross validation (CV) approach where all epochs from one subject were left out for each fold. For comparison we also implemented CV by epoch, where epochs from a single subject can appear in both the training and testing splits. To ensure an equitable comparison, we ran both CV by epoch and CV by subject conditions with the same number of validation folds, the same train-validation split sizes, and for the same number of training iterations. After hyperparameters were manually tuned, model parameters were frozen and testing was performed on the external dataset. Classification metrics from training and testing data were reported both at the subject level (N=subjects) and epoch level (N=epochs). To classify at the subject level, predictions for each of their corresponding epochs were obtained and a single classification was determined using a majority vote.

## Results

### Comparison of Validation Approach

Figure 2 and Table 1, summarize the comparison between CV by epoch and CV by subject. We see that performance is significantly increased when CV is done by epoch which indicates that the information leakage between the training and test data has a dramatic effect on performance metrics. Conversely, our CV by subject achieved rather modest performance when metrics are calculated at the epoch level. To supplement these metrics, Figure 3 and Table 2 report the performance metrics of CV by subject when epochs are aggregated and a classification is made for each subject as a whole. Despite the modest epoch level metrics of CV by subject, we see that the ability to aggregate multiple epochs over an individual patient can help overcome some of the epoch-level heterogeneity and improve performance without compromising real world applicability.

**Figure 1.**
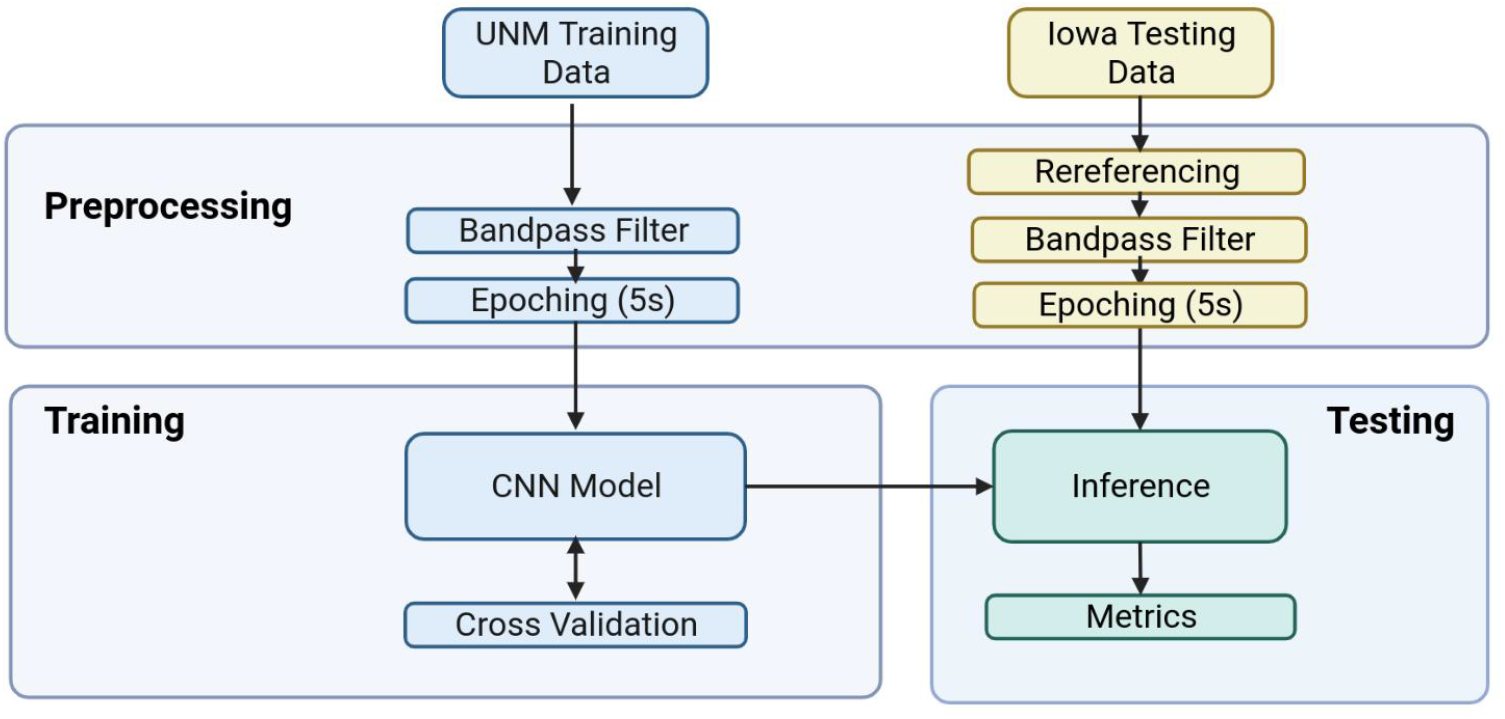
Overview of EEG preprocessing and machine learning pipeline.

**Figure 2.**
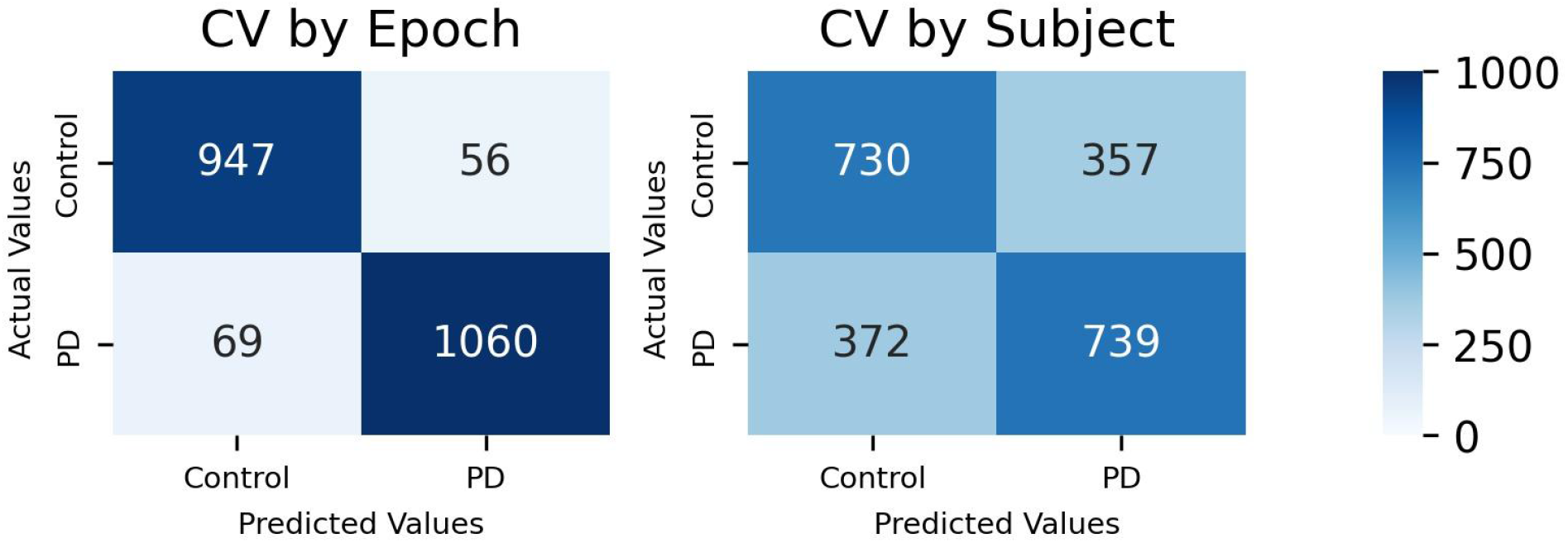
This is a comparison of confusion matrices generated by classifying individual epochs using K-fold cross validation [left] and leave-one-subject-out cv [right].

**Figure 3.**
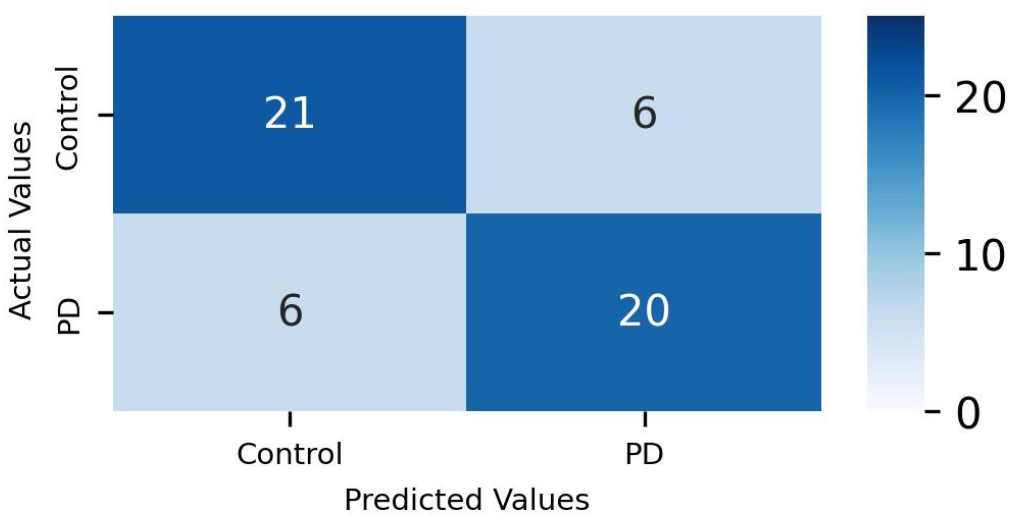
This is a confusion matrix showing the classification of subjects using leave-one-subject-out cross validation.

**Table 1.** This table summarizes the comparison of classification metrics between CV by epoch and CV by subject. This experiment was performed using the training data and classification metrics are reported treating each epoch as a sample.

| Metric (epoch classification) | CV by Epoch | CV by Subject | Difference: |
| --- | --- | --- | --- |
| Accuracy | 0.941 | 0.692 | 0.294 |
| F1 | 0.944 | 0.694 | 0.250 |
| Sensitivity | 0.939 | 0.665 | 0.274 |
| Specificity | 0.944 | 0.722 | 0.222 |

**Table 2.** Summary of classification of training data at the subject level using cross validation by subject. Classification metrics reported treating each subject as a sample.

| Metric | Subject Classification |
| --- | --- |
| Accuracy | 0.774 |
| F1 | 0.769 |
| Sensitivity | 0.769 |
| Specificity | 0.778 |

### External testing

The most suitable metric to evaluate real-world performance of a predictive model is to test it on completely independant data ideally collected at a different site by separate investigators. Here we show how our model performs on the UI test set both in epoch and subject classification tasks in Figures 4-5 and Figures 6-7, respectively. Indeed, regardless of the training approach you use, the testing metrics shown in Table 3 are much closer to the conservative estimates provided by subject-level holdout as compared to estimates using cross validation experiments where subjects’ epochs are intermixed within training and validation sets. Encouragingly, Figure 6 and Table 3 show that high accuracies can be achieved when analyses are conducted on the subject level allowing potentially non-informative epochs to be averaged out and diluted across the entire EEG recording session. To further understand the performance of our model, we plotted the distribution of classification success by subject (Figure 7). In 3 of the 4 errors, there seems to be strong agreement of the intra-patient epochs.

**Figure 4.**
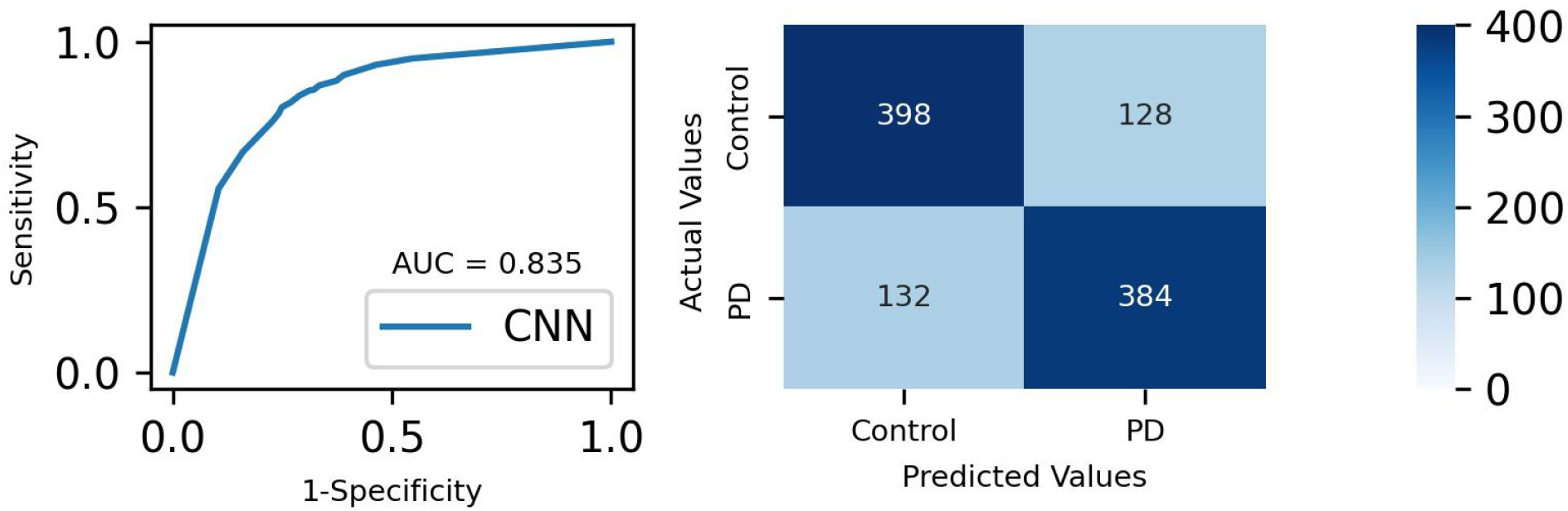
This figure is a sample receiver operator characteristic curve [left] and confusion matrix [right] describing the performance of the CNN classifier on the external UI test-set. Confusion matrix is calculated using each epoch as a single sample.

**Figure 5.**
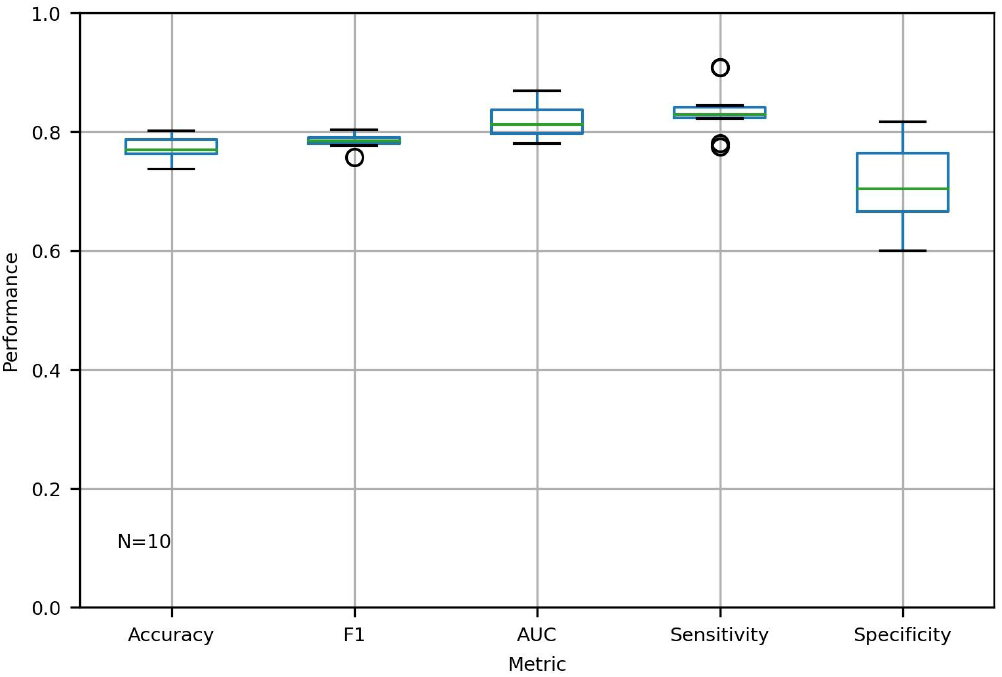
This is a boxplot showing the distribution of test dataset classification metrics calculated for ten replicates of our CNN model classifying individual epochs.

**Figure 6.**
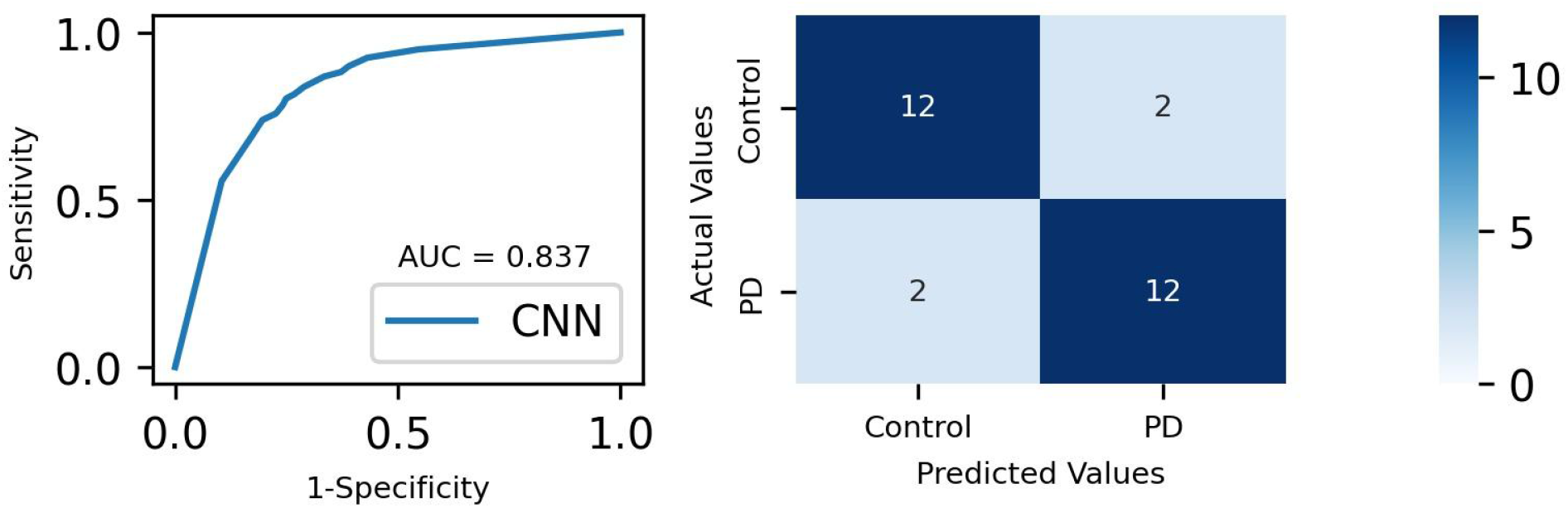
This figure is a sample receiver operator characteristic curve [left] and confusion matrix [right] describing classification of the test set. Metrics were generated using each subject as a sample.

**Figure 7.**
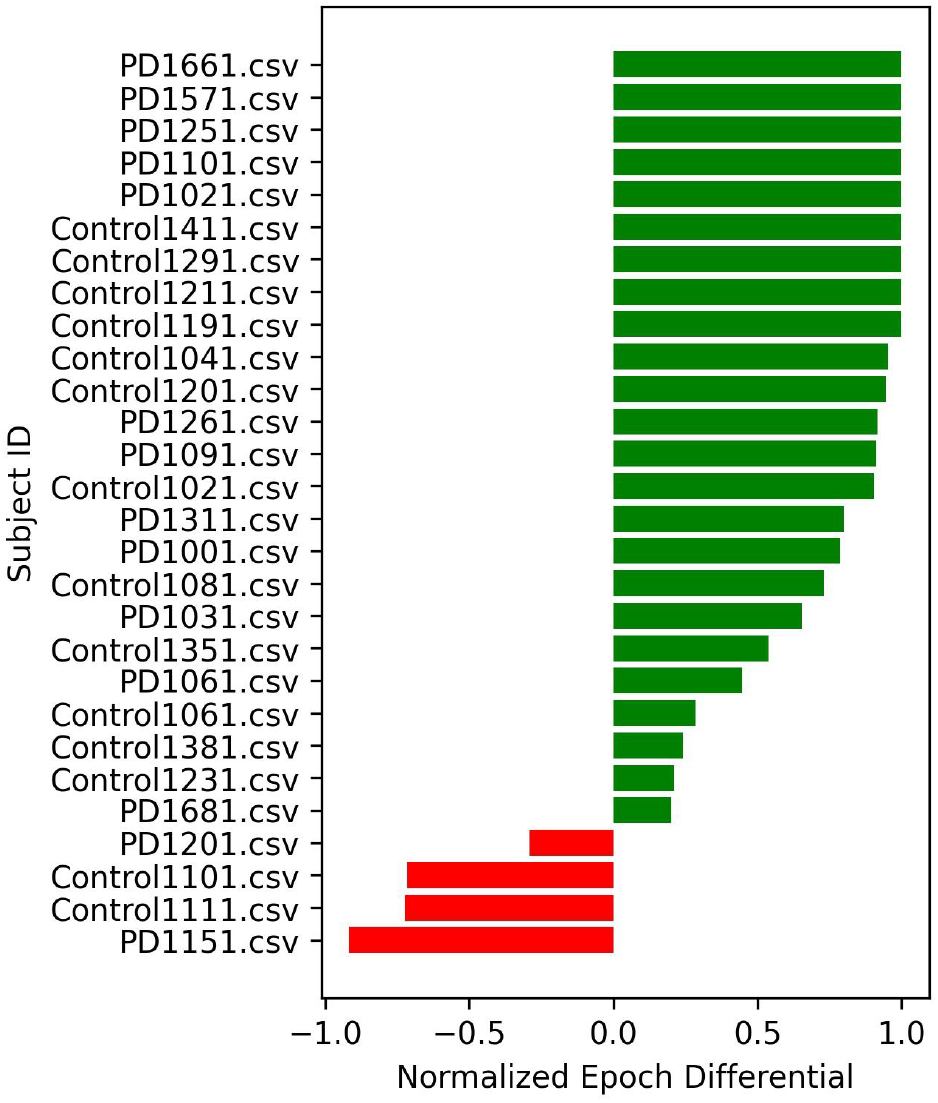
This is a bar graph showing the normalized differential between correct and incorrect epochs, for each subject. Positive scores (green bars) indicate that the subject was classified correctly; negative (red) indicates an incorrect subject classification. A score of one means that all epochs were classified the same way, while a score of zero indicates equal classifications in either class.

**Table 3.** Summary of the mean performance of ten CNN replicates classifying the external UI testing data. Metrics are calculated treating each epoch as a single sample [center] and each subject as a sample [right].

| Metric (by epoch) | Mean Score (Epoch Classification) | Mean Score (Subject Classification) |
| --- | --- | --- |
| Accuracy | 0.772 | 0.838 |
| Sensitivity | 0.835 | 0.886 |
| Specificity | 0.710 | 0.790 |
| F1 | 0.784 | 0.847 |
| AUC | 0.817 | 0.889 |

## Discussion

We have presented a conservative and intuitive CNN-based pipeline to achieve stable and more predictable classification of PD from EEG data. To the best of our knowledge, this work is the first to demonstrate a stable DL classification ability on resting-state EEG data from patients with PD as shown by the external test set. Importantly, we reported a diversity of classification metrics and ROC curve showing that our CNN model accomplishes balanced classification and is not driven by confounders or training artifacts. While it provides more conservative estimates of performance, our approach to CV may be critical to understanding the potential of CNN classifiers when external datasets are not available. Furthermore, our code is made publicly available to demonstrate the reproducibility of our results and to help others prevent limitations of reporting performance metrics that could be contaminated by information leakage between training and test epochs.

EEG provides a non-invasive tool to explore brain activity across diverse disease types, but large cohorts have thus far been limited. One nearly universal practice in the field of EEG to overcome this limitation is to divide subjects’ data into shorter epochs of data. This reduces the number of input features while also increasing the number of unique samples available. Another technique that has been common in deep learning classification of PD-EEG data is to use K-fold cross validation to show the robust classification ability. However, Aljalal et al. noted that the combination of epoching and cross validation leads to an information leakage between the train and test sets since both sets will include epochs from the same subjects [6]. To highlight the effect of this information leakage, we performed an investigation using the previous approaches, and found that classification metrics may be dramatically exaggerated (by 20% or more) to values that are not representative of the classifier’s true performance on the test set.

To generate more representative predictions, our pipeline first partitions cross validation folds by subject, then proceeds to segment data into epochs. This achieved accuracies of 69% and 77% corresponding to classification performed on each epoch and subject respectively. The later metric is perhaps the most relevant for real-world deployment and use. Furthermore, testing was performed on completely unseen subjects’ data collected at a different site achieving an epoch accuracy of 77% and a subject accuracy of 84%. While this approach may lead to classification metrics that are more modest than others reported in the literature [7, 10, 14], it may provide a more conservative and robust prediction of how these classifiers are anticipated to perform in real world settings. For future work, we recommend that cross validation performed at the subject level (i.e. leave-one-subject-out) remains a standard to prevent these limitations.

While this finding that information leakage can improve performance was expected, it may still provide valuable insight. For example, this may be due to a subject-specific biosignature present in the EEG brainwaves. This suggests that longitudinal monitoring of these subject-specific signals using medical-grade (or potentially even lower cost EEG devices) could provide personalized biomarkers for highly complex and heterogeneous diseases that do not show reliable population level changes. Alternatively, the classifier may be overfitting on the specific experimental conditions of the recordings (humidity, time of day, subject mood etc.). Further research is required to understand what features of the data are being used by the models to overfit.

We found that for three of the four misclassified subjects, there was strong intra-subject agreement of epochs (Figure 7). This may suggest that there may be some overlapping EEG signals in some patients that cannot be resolved using population level EEG patterns alone. It is possible that this heterogeneity of PD could be modeled with either more training data available or with more complex models that can better account for the variation. Future work to improve performance includes performing a search for more effective models, preprocessing techniques, and epoch aggregation methods.

## Conclusion

Here we demonstrated the ability of a DL classification workflow to generalize on an unseen testing dataset collected by different investigators at a separate site than the training dataset. Our results using such a simple workflow and model offer a more valuable standard for benchmarking future techniques and disease types. These results hold promise that improvements may be made by using more sophisticated DL approaches.

## Online Content

Our code and the pre-processed data used in these experiments can be accessed at https://github.com/diamandis-lab/CNN_EEG_PD

## Data Availability

Our code and the pre-processed data used in these experiments can be accessed at
https://github.com/diamandis-lab/CNN_EEG_PD

https://github.com/diamandis-lab/CNN_EEG_PD

## Abbreviations

CNN: Convolutional neural network
CV: cross validation
DL: deep learning
EEG: Electroencephalography
PD: Parkinson’s Disease
UI: University of Iowa
UNM: University of New Mexico

## Ethics Approval

EEG data used in this study were retrieved from previously published, anonymized, publicly available resources; thus, additional institutional research ethics board approval was not applicable.

## Acknowledgements

R. S. is supported by a NSERC Canada Graduate Scholarship (Master’s), and the University Of Toronto Fellowship - Medical Biophysics. This work and the Diamandis Lab are supported by an Adam Coules Research Grant. No sponsors had any role in the study design and execution or in the process of writing and submitting the manuscript.

## Author Contributions

R.S. performed analysis and generated figures. R.S. and P.D. designed experiments and wrote the manuscript. P.D. Supervised the work.

